# Cost-effectiveness modelling to optimise active screening strategy for *gambiense* human African trypanosomiasis in the Democratic Republic of Congo

**DOI:** 10.1101/2020.08.03.20167296

**Authors:** Christopher N. Davis, Kat S. Rock, Marina Antillón, Erick Mwamba Miaka, Matt J. Keeling

## Abstract

*Gambiense* human African trypanosomiasis (gHAT) has been brought under control recently with village-based active screening playing a major role in case reduction. In the eve of elimination, we investigate how to optimise active screening in villages in the Democratic Republic of Congo, such that the expenses of screening programmes can be efficiently allocated while continuing to avert morbidity and mortality. We implement a cost-effectiveness analysis using a stochastic gHAT infection model for a range of active screening strategies and we calculate the net monetary benefit (NMB) of each strategy. High-coverage active screening strategies, occurring approximately annually, attain the highest NMB. We find that, for strategies stopping after one to three years of zero case reporting, the expected cost-benefits are very similar and we highlight the current recommended strategy (three years before stopping) is likely cost-effective, in addition to providing valuable information on whether transmission has been interrupted.

## Introduction

Despite the continued decline in the annual number of reported cases of *gambiense* human African trypanosomiasis (gHAT), accounting for less than 1,000 new cases reported in 2019 [1], the disease persists in many of the historically endemic sites in Western and Central Africa. This vector-borne disease, transmitted by a bite from a tsetse infected with the parasite *Trypanosoma brucei gambiense*, is typically — although not always — fatal when untreated [2]. Human African trypanosomiasis (HAT), which includes both *gambiense* and *rhodesiense* forms, caused an estimated 1,364 deaths in 2017 and approximately 78,990 disability-adjusted life years (DALYS) [3]. The disease has been targeted for elimination by the World Health Organization (WHO); first, for elimination as a public health problem by 2020 and then for elimination of transmission (EOT) by 2030 [4, 1]. To achieve these targets, there are several recommended strategies to reduce the transmission and burden of the infection, which are constituted primarily of the medical interventions of active screening and passive surveillance.

Passive surveillance depends on the ability of fixed health centres to test for the infection and carry out treatment on self-presenting individuals, typically upon the onset of symptoms [5]. Screening and treating infected individuals both allows the infected people to be saved from a potentially fatal disease, but it also prevents further spread of infection via tsetse.

Traditionally, the most effective form of controlling gHAT infection, however, has been active screening and treatment [6, 7, 8]. Active screening is carried out by mobile teams that travel to villages in focal disease regions and target the screening of the whole population for gHAT; those determined to have the infection can then be treated at the closest health facility offering treatment. The initial screening test is typically a serological test for the presence of the antibody called the Card Agglutination Test for Trypanosomiasis (CATT) [9], although recently rapid diagnostic tests (RDTs) have also been utilised as an initial diagnostic [10, 11, 12]. Confirmation of the infection is then carried out by viewing the parasite under microscopic examination; traditionally this was followed by staging of the disease, which consists of a lumbar puncture to determine whether the parasite has infected the central nervous system — considered the second stage of disease [13]. However, the recently approved drug, fexinidazole [14], should remove the need for lumbar puncture in most cases although retaining the requirement of parasitological confirmation [15].

Active screening has been very effective in reducing case numbers and still plays an important role in maintaining surveillance and treatments where access is problematic, yet it is an expensive intervention in terms of both time and money [16, 10]. As local elimination of gHAT occurs in focal areas, active screening will likely be scaled back and gHAT testing will become better integrated into fixed health facilities, as resources can be reallocated and it becomes unnecessary to screen entire village populations for the infection [17]. In this situation, reactive screening can be implemented, whereby after a number of successive active screenings in which no cases are detected, the screening stops unless a new case is passively reported, upon which a ‘reactive’ screen would occur [11]. Several active screening strategies have been proposed, including a recommendation of three repeated screening rounds with one-year [18] or six-month intervals [19]. WHO guidelines currently recommend annual screening for three years of zero case reporting before stopping in previously endemic villages [4].

Mathematical models of gHAT have been used for the prediction of future case numbers and evaluation of a range of plausible control strategies [20, 21, 22, 23, 24, 25, 26, 27, 28]. However, these have typically considered the infection dynamics and the impact of interventions without accounting for the costs of implementing such strategies. Here, we explicitly use a stochastic model of gHAT infection in a village population, developed in Davis et al (2019) [25], to simulate different plausible active screening programmes alongside passive surveillance, allowing us to quantify the relative costs of implementation as well as the health effects compared to a baseline of passive surveillance (the comparator strategy). We use parameters matched to screening and incidence data from the health zone Kwamouth, in Mai-Ndombe province of the Democratic Republic of Congo (DRC) (formerly in Bandundu province). Kwamouth is in a historically high-endemicity gHAT area of the DRC, the country that contributes 70% of all global gHAT cases in 2019 [1]. We also present results from a moderate-endemicity health zone, Mosango, in Appendex 3.

The costs of gHAT interventions have been previously been evaluated [29, 30, 10, 31, 32] and the different strategies have been considered for large populations [33, 34]. We consider the effect of active screening on individual villages in the drive for EOT, by determining how active screening can be best implemented to achieve this goal whilst providing value for money.

## Results

### Breakdown of costs of active screening

We use the stochastic compartmental model from Davis et al (2019) [25] to simulate different strategies for active screening that vary: screening coverage *c*, screening interval *t*, active zero-detections *z*_*a*_, and reactive zero-detections *z*_*r*_ (see Table 1). In the model, individuals in the human population are classified as either high-risk or low-risk [21], whereby the high-risk population — a small minority, which has been previously estimated to be 9.8% in the study health zone of Kwamouth [35] — have a higher exposure to tsetse and do not participate in active screening. This means that the screening coverage is assumed to have a maximum of 90%, since only the low-risk human population participate (randomly) in active screening. Reactive screening is a resumption of active screening and occurs upon detection of a case in passive surveillance after active screening has been stopped (see Appendix 4).

**Table 1:** Descriptions of the variables used for defining an active screening strategy.

| Variable | Name | Definition | Value range |
| --- | --- | --- | --- |
| $c$ | Screening coverage | Proportion of the village population screened in a visit. | 0–90% |
| $t$ | Screening interval | Time between active screening visits to a village. | 0.25–5 years |
| $z_a$ | Active zero-detections | Number of consecutive active screenings where no cases are detected for the cessation of active screening. | 1–5 screenings |
| $z_r$ | Reactive zero-detections | Number of consecutive reactive screenings where no cases are detected for the cessation of reactive screening. | 1–3 screenings |

The cost of an active screening strategy is a function of several component costs: implementing the screening test, confirmation of the infection, carrying out treatments, setting up and maintaining the mobile screening teams. Moreover, active screening may impact the number of passive tests and treatments performed. In the current work, we do not consider the additional costs of passive surveillance, such as capital costs, only the costs directly affected by active screening. The costs of active screening strategies will vary depending on the type of screening test and treatment used, and also the type of mobile screening team; while a traditional truck team that can carry more tests and equipment, such as a generator, a motorbike team that can reach more remote villages [36, 31].

As well as considering the changes in monetary costs, we want to consider the change in the health benefit of implementing different active screening programmes; therefore, we consider the number of DALYs averted [37]. The number of DALYs are the discounted sum of the number of years of life lost (YLL) and years lived with disability

(YLD), where YLL is the number of years of life lost due to premature death and YLD is the number of years of healthy years lost with a weighting for the severity of the condition [38]. We calculate the number of DALYs averted by a particular screening strategy against a comparator consisting of passive surveillance and no active screening.

We evaluate the net monetary benefit (NMB) to assess the cost-effectiveness using a 30-year time-horizon. For each active screening strategy, the net monetary benefit (NMB) was calculated as:

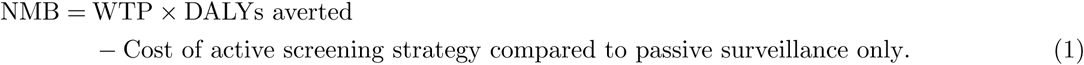

The willingness to pay (WTP) is the maximum amount of money that the funder is prepared (willing) to pay to avert one DALY. The number of DALYs averted is the change in DALYs between a strategy and the comparator (specifically, implementing only passive surveillance but no active screening). The change in costs is the cost of implementing the active screening strategy, including the consequential change in cost of passive surveillance, minus the cost of baseline passive surveillance. A particular strategy is more beneficial than the comparator strategy if the NMB is positive, conditional on a WTP value. In the context of uncertainty (repeated draws of the simulation) the optimal strategy will have the highest mean NMB. Because the NMB is so highly dependent on the value of the WTP, we consider a range of fixed WTP thresholds, such that decision-makers can heed recommendations according to the typical cost-effective thresholds in their programs.

The correct WTP is the cause of much debate. Typically, the WTP is taken as the product of the gross domestic product (GDP) per capita of a country and a multiplying factor. This factor, *WTP*_*c*_, is traditionally given as 3 [39], but this is often considered too high for low-income countries [40] and so 0.5 is also used [41, 42]. We note that in the context of elimination, a funder may be willing to pay more for the additional benefit of reducing the number of infections to zero, but we simply leave this choice to the funder. In addition to the WTP, the NMB will be affected by the population size of a village, *N*_*H*_, the proportion of infections that go undetected by active screening but that are detected and treated passively *p*_*t*_, and the initial level of infection in the population.

We first consider an active screening strategy with a typical screening coverage of 55% (see Appendix 3), carried out annually, and with three active zero-detections and one reactive zero-detection required for the cessation of screening (*c* = 55%, *t* = 1 year, *z*_*a*_ = 3, and *z*_*r*_ = 1). We assume a village population of size *N*_*H*_ = 1, 000 starting from endemic equilibrium (as determined from the deterministic version of the model), and with 27% of infections undetected in active screening treated passively (*p*_*t*_ = 27%). We calculate mean values of one million stochastic realisations of the process. For this strategy, the prevalence in both the human and tsetse populations rapidly decays towards zero (Figure 1A). The annual cost of implementing this strategy also decreases with time (Figure 1B); this is in part due to 3% discounting, the method of adjusting future costs to present-day values (which is applied to both costs and DALYs averted) [43], but also because decreasing the prevalence of infections in the population reduces the required number of treatments. Even with no active screening, infections may die out in the village due to ‘stochastic fade out’, when local disease extinction occurs purely by chance. There is a small annual increase in costs after twelve years, since the difference in the number of infections treated in passive surveillance is smaller in later years. However, costs decay towards zero as the probability of gHAT extinction increases with time; while some recurrent costs will remain, the number of treatments will decline in time with the corresponding reduction in infections. In addition, the costs also decrease when the consecutive zero-detection threshold is reached, as the cost of the active screening is completely removed. With the assumption that *p*_*t*_ = 27% and *c* = 55%, the number of DALYs averted will initially increase each year because more people are treated after detection during active screening. However, in later years, DALYs averted will decline due to fewer infections under either strategy, implying that the differential impact in the active screening strategy (versus passive surveillance alone) is most substantial in the early years of implementation because both the strategies and passive surveillance are expected to lead to elimination, albeit at a different speed (Figure 1B).

**Figure 1:**
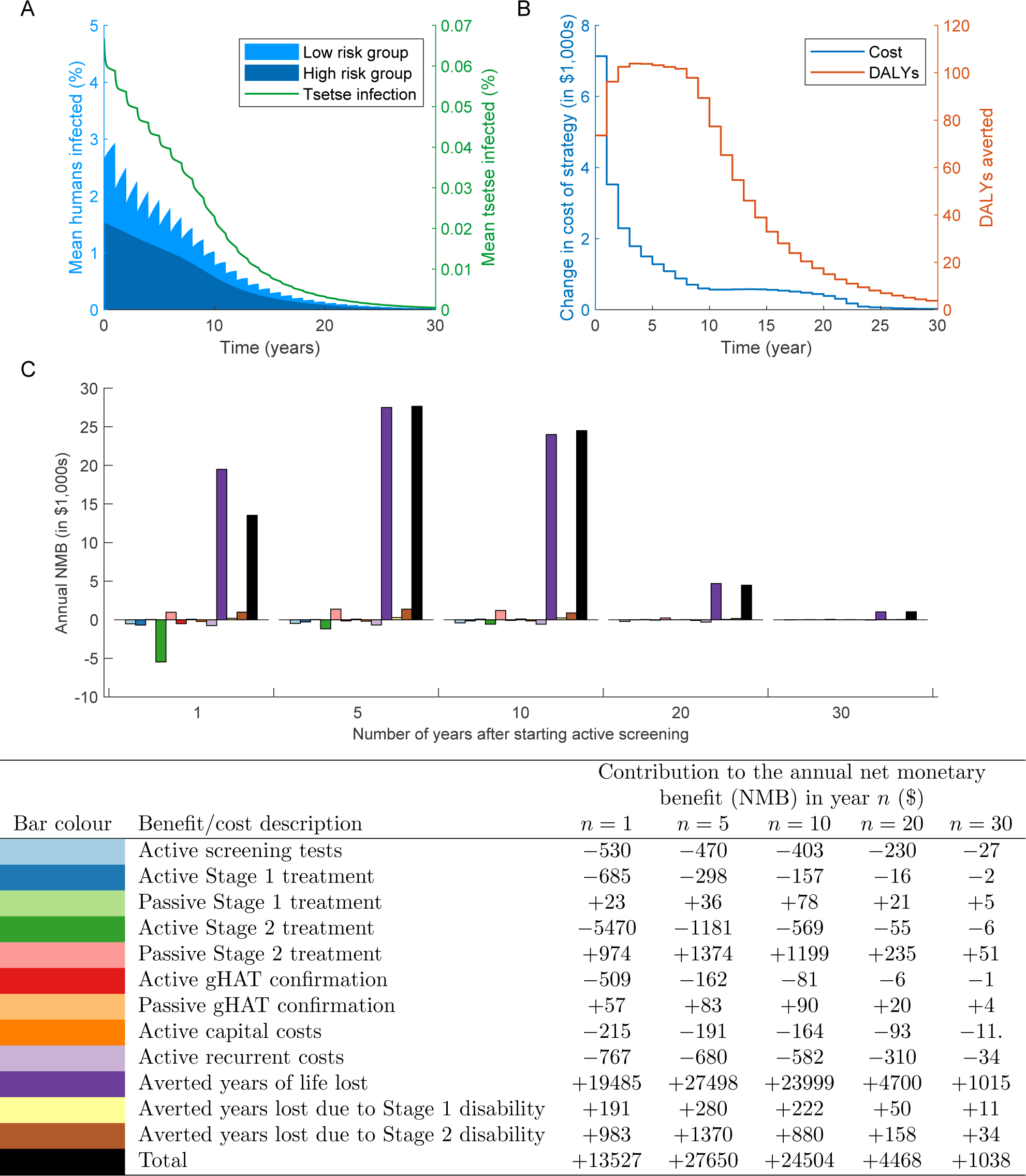
The cost of active screening for a coverage of 55%, a screening interval of 1 year, stopping active screening after 3 screenings when no cases are detected and stopping reactive screening after 1 screening with no cases, under the assumption of *WTP*_*c*_ = 0.5. Mean values for all quantities are take from one million stochastic simulations. (A) The number of infected people dramatically decreases with time for this coverage (total shaded blue area, with left axis) with the majority of these infections being in the high-risk group (darker blue fraction). The proportion of tsetse that are also infective is reduced with time (green line, with right axis). (B) The total change in costs of implementing a particular screening strategy (left axis) and the number of DALYs averted from the baseline of only passive surveillance (right axis). (C) The contribution to the cost from each component of the cost function for years 1, 5, 10, 20 and 30 after starting an active screening program. Full costs are given in the table in the bottom row of the table. A population size of *N*_*H*_ = 1, 000 is used. All costs are denominated in 2018 US dollars.

A breakdown of the components of the NMB of implementing this active screening strategy shows that the biggest costs are the treatment from active screening, the recurrent costs of the active screening and screening populations with the CATT test (Figure 1C). However, assuming a WTP of 50% of the GDP per capita of the DRC (*WTP*_*c*_ = 0.5 giving WTP equal to $280.89 [44]) the monetary benefit in reducing the years of life lost is dominant and the biggest factor in maximising the NMB. The total NMB (black bars) shows the full benefit of this active screening strategy is always positive with *WTP*_*c*_ = 0.5 and so, on average, this strategy is better than the comparator of no active screening. Further into the future, the NMB moves closer to zero, both because of discounting and because there is a higher probability the infection will be locally extinct, and so active screening not required. Note that the NMB of passive surveillance is positive because the introduction of active screening and treatment means that fewer passive confirmations and treatments will need be carried out, reducing the cost. The table in Figure 1C shows the NMB breakdown in full.

### Drivers of net monetary benefit across strategies

We performed a four-way sensitivity analysis of the NMB for the WTP, the treatment coverage in passive surveillance *p*_*t*_, the screening coverage *c* and screening interval *t* (Figure 2). We considered the mean value of the NMB for one million simulations for every screening strategy considering screening coverage *c* and screening interval *t* for a large number of values (stated in Methods and Materials), but discretising WTP and *p*_*t*_ to three and two values respectively for the presentation here. To view how the exact optima change with these model parameters and all the specified values for the cost and benefit parameters (including, treating WTP and *p*_*t*_ as continuous variables) visit the supplementary R Shiny [45] app at https://christopherdavis.shinyapps.io/optimising-ghat-active-screening/. This app allows the sensitivity of the optimal solution to be found on any updated costings for the active screening strategies.

**Figure 2:**
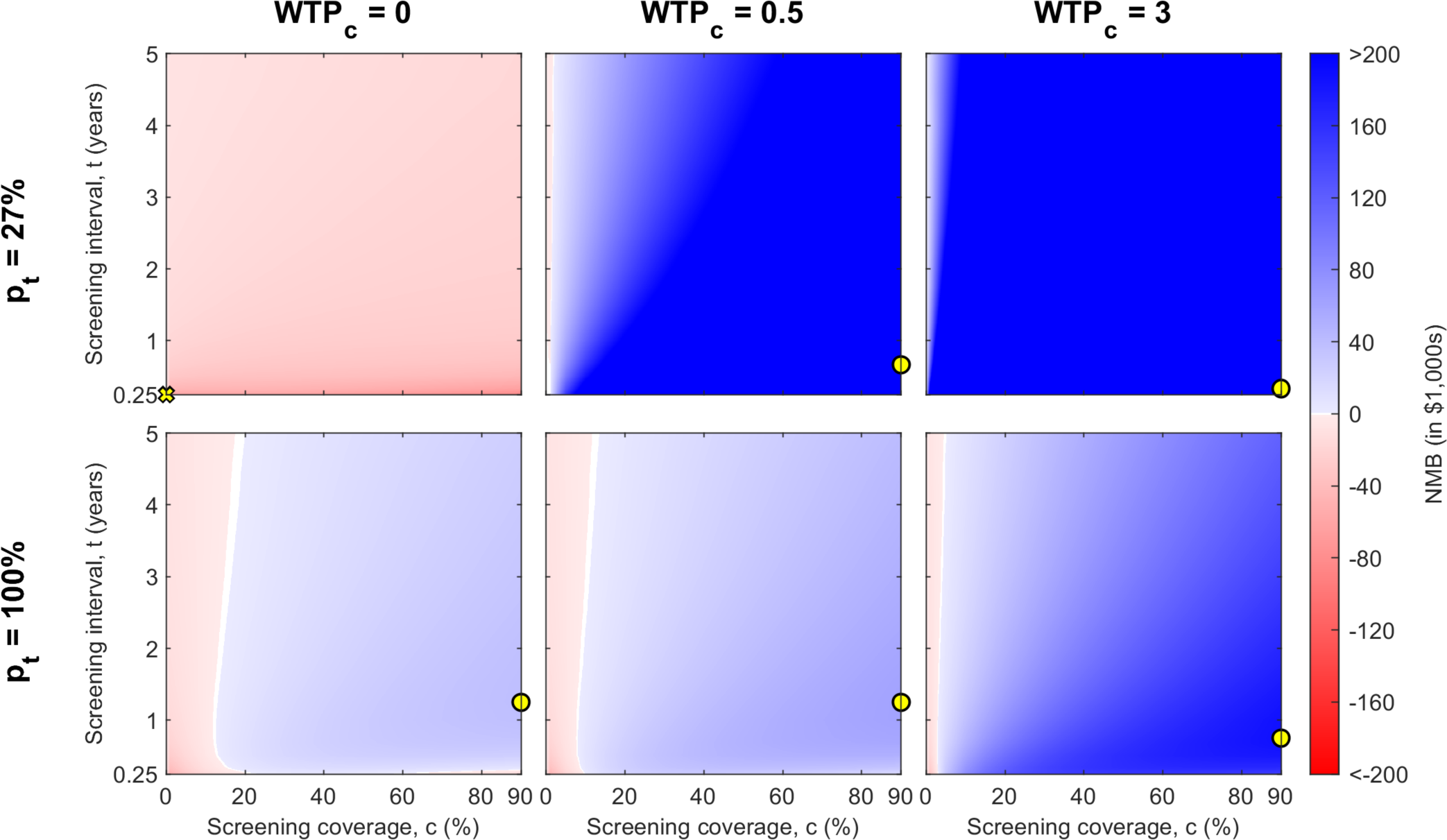
The mean NMB of different active screening strategies for given WTP per DALY averted (given as multiplication factor *WTP*_*c*_ of GDP per capita from the DRC) and the proportion of passive infections that are treated, *p*_*t*_. The red areas show a negative NMB, while blue areas are positive NMB, with white at the boundary of no change. The maximum NMB for each *WTP*_*c*_ and *p*_*t*_ combination is marked by a yellow circle on each heatmap, with a cross if the maximum is for no active screening (only observed here for *WTP*_*c*_ = 0 and *p*_*t*_ = 27%). A population size of *N*_*H*_ = 1, 000 is used and we fix *z*_*a*_ = 3, the number of consecutive active screening rounds with zero-detections necessary to cease operations.

### The interaction between WTP and *p*_*t*_

The NMB for some strategies is highly dependent on both the WTP and the proportion of passive infections treated (*p*_*t*_), with (1 *- p*_*t*_) suffering disease-induced mortality, but the impacts of these factors on NMB are neither linear nor consistent. For instance, at low values of *p*_*t*_ (*p*_*t*_ = 27%), the adoption of active screening strategies (of any frequency or coverage) is substantially different in the WTP value range of 0–0.5, but increasingly high WTP values would not yield different strategies. However, at high values of *p*_*t*_ (*p*_*t*_ = 100%), the adoption of active screening strategies at any frequency or coverage level are incumbent on very high WTP values.

### The impact of WTP and *p* _*t*_ on optimal values of screening coverage and interval

When assumptions about WTP and *p*_*t*_ are fixed, the screening coverage has a greater impact on whether the NMB is positive than the screening interval. Low screening coverage levels (¡20%) can be insufficient to obtain a positive NMB, while the screening interval does not change the sign of the NMB for most coverage levels, unless the interval is very small (0.25 years). The question of what value to fix for WTP and *p*_*t*_ has important implications on the optimal choice of screening coverage and optimal screening interval.

First, the optimum screening interval will be approximately one year under all assumptions of passive surveillance treatment coverage (*p*_*t*_) and WTP (yellow dots in all panels of Figure 2). For *WTP*_*c*_ = 0.5 and *p*_*t*_ = 27% (our standard assumption), the maximum mean NMB is found when the screening coverage is 90% and the screening interval is 0.67 years (Figure 2; the yellow dot in top centre panel). For *WTP*_*c*_ = 0.5 and *p*_*t*_ = 100%, the maximum mean NMB is also at the maximum screening coverage, but the higher treatment coverage (*p*_*t*_) indicates that the optimal screening interval is of 1.25 years. The optimal screening interval the same for all values of the number of zero-detections, but the minimum NMB was found at *z*_*a*_ = 1 and *z*_*r*_ = 1 (see https://christopherdavis.shinyapps.io/optimising-ghat-active-screening/). It is notable that a very high WTP (3 times the GDP per capita) lends strong support for shorter screening intervals, favouring screenings in a village multiple times a year.

Second, and turning the attention from the active screening interval to the coverage, we found that in terms of NMB the screening coverage has an inverse relationship with the treatment coverage under passive surveillance (*p*_*t*_). When *p*_*t*_ = 100%, the assumption is that all infections are eventually treated, implying no loss of life, which would otherwise be a large component of the change in NMB. Thus, under the assumption of a high *p*_*t*_, active screening strategies with low coverage will be preferable, indicating that high costs of screening implementation will hardly be justified by DALYs averted, as passive surveillance is already very effective (Figure 2). In contrast, under an assumption of a lower *p*_*t*_, high active screening coverage is needed to compensate for lower treatment coverage in passive surveillance.

### Sensitivity analysis of village characteristics and maximum net monetary benefits

We present the active screening strategy that gives the maximum mean NMB for a range of WTP values, examining the role of the treatment coverage under passive surveillance, the village population size, and the endemicity status of the village. To do this, we apply costs (see Appendix 2) to the mean simulation outputs to see which strategy provides the largest NMB. Each line in Figure 3A–C shows the value of *c, t* and *z*_*a*_ that together give the optimal strategy for given values of *p*_*t*_ and WTP. For instance, assuming that *WTP*_*c*_ = 0.5 and *p*_*t*_ = 27%, the maximum NMB is obtained for *c* = 90%, *t* = 0.75 years, and *z*_*a*_ = 1 (Figure 3A–C). Figure 3D–F considers the same results for villages of different sizes (*N*_*H*_) and with different initial conditions of the simulation (endemic, disease-free, and endemic with importations), all with *p*_*t*_ = 27%. We fix *z*_*r*_ = 1 for all simulations.

**Figure 3:**
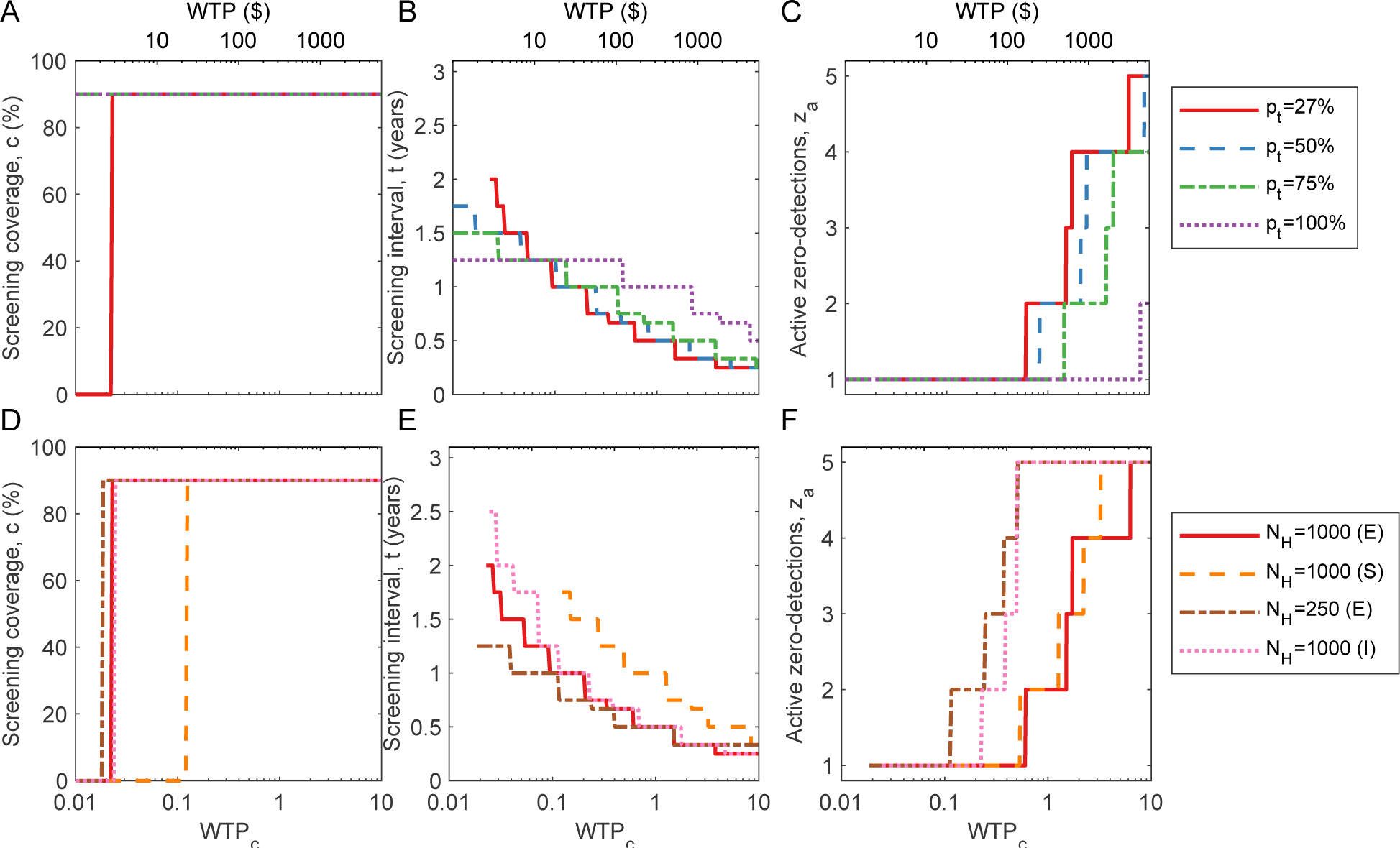
Theoretical optimum strategy for the mean simulation of infection dynamics given a range of WTP values (horizontal axis). (A)–(C) examine the impact of different treatment coverage (*p*_*t*_) on (A) the optimal screening coverage, (B) the optimal screening interval and (C) the optimal number of zero-detections required to stop screening, to achieve the highest NMB for given WTP. These results assume a village population of 1,000 where the disease in endemic. (D)–(F) examine the impact of different assumptions about population size and endemicity on (D) the optimal screening coverage, (E) the optimal screening interval and (F) the optimal number of active zero-detections required to stop screening. The demography and disease endemicity assumptions are as follows: a population of population 1,000 where the disease is endemic (‘1000 (E)’), a village of population 1,000 where only one person is initially infected (single infection, or ‘1000 (S)’), a village of population 250 where disease is endemic (‘250 (E)’), and a village of population 1,000 where disease is endemic and there exists a small probability of infectious importations (‘1000 (I)’). We fix *z*_*r*_ = 1 for all simulations.

Screening coverage, *c*, is optimal at 0% (not doing any screening) or at a very high coverage (the maximum of 90%) (Figure 3A). For *p*_*t*_ = 27%, active screening at the maximum coverage is optimal for *WTP*_*c*_ > 0.02 (Figure 3A). This means if there is no WTP to avert DALYs (the decision-maker wants to remain cost-neutral), it is best not to incur any screening costs, since the NMB will be negative, however, if the WTP is above threshold 0.02 of GDP per capita, it is optimal to screen entire village populations to reduce the prevalence and prevent further transmission. The threshold WTP where maximum screening coverage is optimal is also influenced by *p*_*t*_; the WTP threshold decreases for larger values of *p*_*t*_ (above 27%) and active screening campaigns are always optimal for higher *p*_*t*_ values. Therefore, at high *p*_*t*_ values (treatment coverage), the additional costs of active screening are justified in order to shorten disease duration and expedite elimination (recovering screening costs via averted treatment costs). The optimal screening interval, *t*, and the number of active zero-detection before screening cessation are more sensitive to the WTP, but have similar patterns across values of treatment coverage (*p*_*t*_).

The optimal screening interval, *t*, decreases with increasing WTP, since DALYs are valued more highly and more frequent screening averts more DALYs (Figure 3B). We also show that for any value of *p*_*t*_, the screening interval must be two years or shorter, typically approximately annually. For most WTP values, a single active zero-detection is enough to justify cessation of active screening, but we note that if the funder is willing to pay more, there is a benefit in repeated active zero-detection campaigns before cessation in order to ensure no resurgence of transmission (Figure 3C). To consider the full range of *p*_*t*_ values a heatmap of the change in cost with respect to both *p*_*t*_ and WTP is given in Appendix 5 Figure 2.

Additionally, we have considered three other scenarios: a smaller village population *N*_*H*_ = 250 at endemic equilibrium; a disease-free population starting with a single infection rather than endemic equilibrium, therefore mimicking a local post-elimination reintroduction of the infection; and a population with a small chance of an imported infection is present (Figure 3D–F). For scenarios where the population size *N*_*H*_ = 1, 000, the results are qualitatively similar: lower active screening coverage is optimal when there are fewer infections (due to the single reintroduction). For a smaller population *N*_*H*_ = 250, it is more effective to have a shorter screening interval and more campaigns that yield active zero-detections to ensure local elimination, since the cost of active screening is smaller when there are fewer people to screen. When there is a small rate of importation of infection, the higher probability of sustaining local infection means that a higher number of active zero-detections *z*_*a*_ are optimal for any given WTP, and therefore active screening must continue for a longer period of time.

### Limiting analysis to practical strategies

According to other literature, a high coverage in active screening with a minimum number of visits is desirable [46], which agrees with our results (Figure 3). However, unlike the screening interval and the number of active zero-detections before cessation, which can be designated by district managers, it is not always possible to achieve a desired screening coverage by either decree or investment because screening coverage depends on the availability and consent of the population [47]. In fact, attendance at active screening is often low [48]; for instance, in Kwamouth during 2000–2016, a median screening coverage of 55% was achieved for all village-level active screenings taken from the WHO HAT Atlas [49, 50] (see Appendix 3 Figure 1).

Therefore, since a high screening coverage cannot be guaranteed, we optimise active screening when we have imposed a maximum on the screening coverage consistent with historic (obtainable) coverage in DRC. A higher maximum level of screening coverage allows for a large screening interval and a small number of zero-detections before cessation of active screening. For the very low minimum screening coverage of 5%, the optimum is screening four times a year *t* = 0.25 and five zero-detections to stop (*z*_*a*_ = 5), while for a high coverage, we see the expected result of *t* ≈ 1 year and *z*_*a*_ = 1 (Figure 4). For the median screening coverage in Kwamouth of 55%, the optimal strategy is an active screening every four months, with two active zero-detection required for cessation. A lower screening coverage can be compensated for by an increase in screening frequency thereby reducing the screening interval *t*.

**Figure 4:**
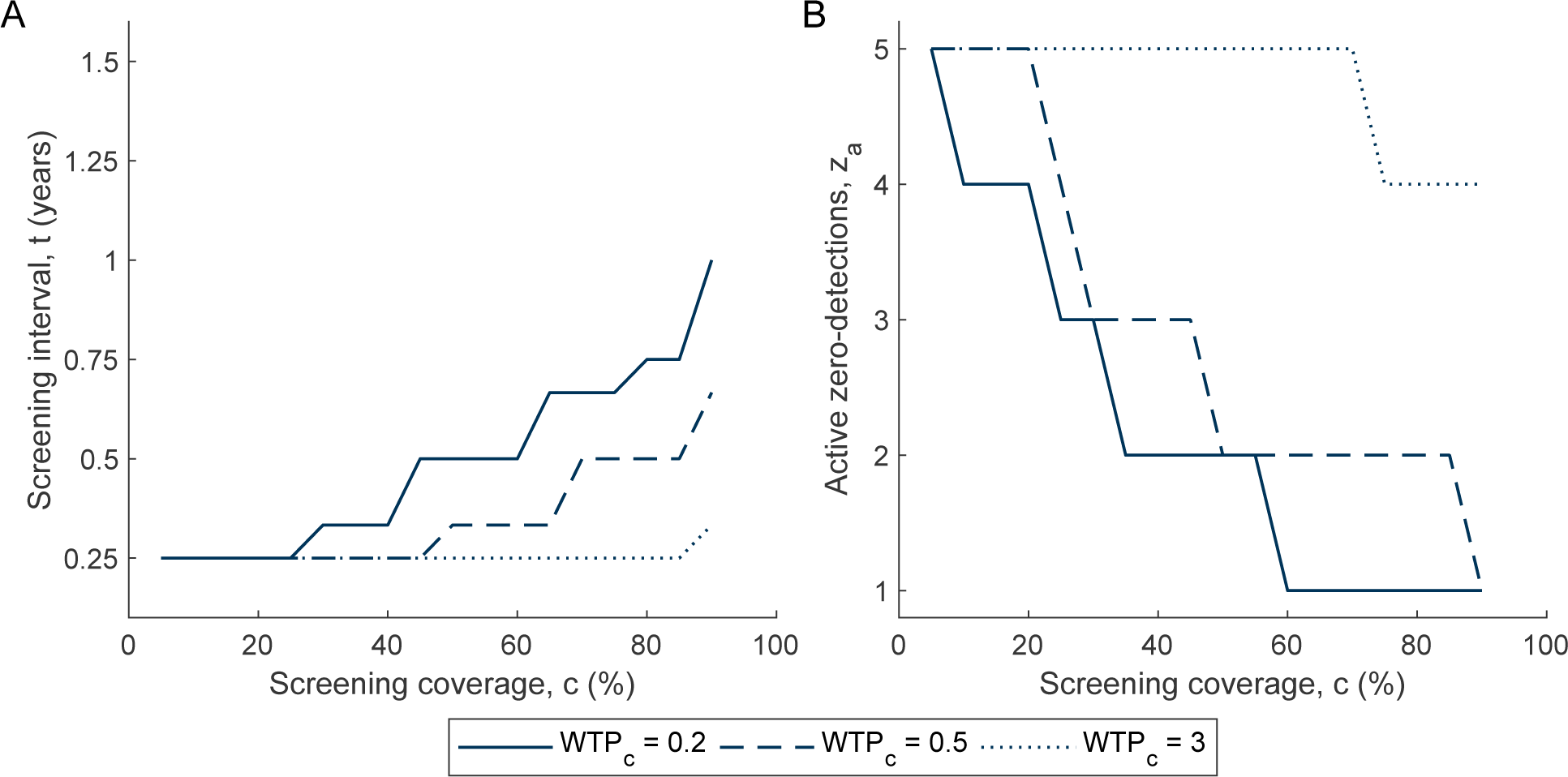
Optimal strategy given a maximum screening coverage informed by historic averages in the DRC. Results are shown for a *WTP*_*c*_ = 0.2, 0.5, 3. *z*_*r*_ = 1, *p*_*t*_ = 27%, *N*_*H*_ = 1, 000 are fixed and the optimum *t* and *z*_*a*_ is found simultaneously. (A) Optimal screening interval *t*. (B) Optimal number of zero-detections to stop screening *z*_*a*_.

### Cost-effectiveness of realistic strategies

While we have determined which strategy, on average, maximises NMB for achievable levels of screening coverage, we now consider the cost-effectiveness of select strategies, restricting the number of strategies under consideration to a smaller number of options. For this process, we have selected seven options: doing no active screening and six realistic proposal schemes for active screening including biennial and annual screening with different cessation criteria. These active screening strategies are shown in Table 2. We assume *z*_*r*_ = 1, *p*_*t*_ = 27% and *N*_*H*_ = 1, 000, and we initialise simulations with conditions consistent with endemic equilibrium.

**Table 2:** Active screening strategies considered in the probability of cost-effectiveness calculations. We show the mean total cost (to nearest dollar) and the total number of DALYs (to the nearest 0.1 DALYs) for each strategy with the 95% prediction intervals across all stochastic realisations. The ACER is the change in cost over the change in DALYs averted as compared to the baseline strategy, while the ICER is compared to the next best strategy (given in the table footnotes). Costs are denominated in 2018 US dollars.

| No. | Strategy | Screening coverage, $c$ (%) | Screening interval, $t$ (years) | Active zero-detections, $z_a$ | Total cost (\$) | Total DALYs | ACER (\$/DALY) | ICER (\$/DALY) |
| --- | --- | --- | --- | --- | --- | --- | --- | --- |
| 1 | Passive surveillance only<sup>1</sup> | 0 | N/A | N/A | 37197<br>[17726, 58928] | 2488.8<br>[1207.8, 3912.6] | Minimum cost | Minimum cost |
| 2 | Biennial screening with one zero for cessation | 55 | 2 | 1 | 55316<br>[33037, 91751] | 1338.9<br>[437.8, 1755.7] | 15.8 | 15.8<sup>2</sup> |
| 3 | Biennial screening with two zeros for cessation | 55 | 2 | 2 | 55862<br>[34302, 91930] | 1339.4<br>[436.3, 1755.5] | 16.2 | Dominated |
| 4 | Biennial screening with three zeros for cessation | 55 | 2 | 3 | 56300<br>[35080, 92357] | 1340.1<br>[434.8, 1756.5] | 16.6 | Dominated |
| 5 | Annual screening with one zero for cessation | 55 | 1 | 1 | 61184<br>[29968, 83446] | 1027.9<br>[570.3, 2262.7] | 16.4 | 18.9<sup>3</sup> |
| 6 | Annual screening with two zeros for cessation | 55 | 1 | 2 | 61871<br>[30847, 83641] | 1027.7<br>[570.6, 2264.3] | 16.9 | 2503.4<sup>4</sup> |
| 7 | Annual screening with three zeros for cessation | 55 | 1 | 3 | 62467<br>[31492, 83773] | 1027.4<br>[572.9, 2261.9] | 17.3 | 2739.0<sup>5</sup> |

<sup>1</sup>The comparator strategy. <sup>2</sup>Relative to Strategy 1. <sup>3</sup>Relative to Strategy 2. <sup>4</sup>Relative to Strategy 5. <sup>5</sup>Relative to Strategy 6.

For our comparator strategy (passive surveillance only) the total cost of implemention is the cost of testing and treating self-presenting individuals infected with gHAT. As previously stated, we do not include the fixed costs of continually operating a passive surveillance network, as we assume that implementing a strategy does not change this cost. Thus, Table 2 shows the average cost of only treating self-presenting patients is $37,197 with 2488.8 DALYs. Since we are considering this our baseline strategy, zero DALYs are averted from this process.

By employing active screening the total costs increase to the benefit of health outcomes; annual screening costs more than biennial screening, but a correspondingly larger number of DALYs are averted under strategies that use annual screening. On the other hand, increasing the number of active zero-detections increases costs but yields few additional DALYs under a regiment of annual screening and even fewer DALYs under biennial screening. Given the 95% prediction intervals for the total DALYs averted for varying just the active zero-detections greatly overlap, there is little basis on which to choose between these strategies other than lowering costs, but the screening interval is much more significant in terms of health benefits conferred (Table 2).

We calculate the ACER as the ratio of the change in cost to change in DALYs averted relative to the comparator strategy, while the ICER is the ratio of the change in cost to change in DALYs averted relative to the next best option (see Table 2). The ICER is the conventional metric for cost-effectiveness: a strategy is cost-effective compared to the next best strategy has ICER ¡ WTP. Active screening at 55% coverage is cost-effective at extremely low WTP values ($15-18 per DALY averted), and the costs-per-DALY of biennial and annual screening strategies are so similar as to suggest that there is little loss in efficiency (few diminishing returns) in undertaking more frequent (yearly) campaigns. Notably, however, a higher WTP ($2,503 per DALY) would be needed to support the choice of strategies with higher values of *z*_*a*_ (multiple active zero-detections), but such strategies should not be implemented at the expense of longer screening intervals. Strategies with higher values of *z*_*a*_ are primarily payment for certainty that transmission chains have been broken, rather purchasing any substantial health burden averted (in terms of DALYs).

### Cost-effectiveness analysis in the presence of parameter uncertainty

Not every village will experience gHAT infection as depicted by the mean infection profile. Hence, we aim to account for uncertainty and present the probability that a strategy is cost-effective. Using the full range of possibilities for the infection dynamics is particularly important for gHAT infection in a village as we know there is potential for large differences between seemingly identical villages, due to the focal nature of the infection [7]. Therefore, we have simulated the infection dynamics of each strategy one million times to compare how the costs and number of DALYs averted can vary.

When there is no active screening, the variations in the cost and DALYs incurred arises from uncertainty in the transmission model (since cost parameters are held constant). By the same token, costs and DALYs are positively correlated because both arise from transmission, passive detection and treatment. The more infections there are in the village, the more treatments will be performed, increasing costs and incurring DALYs. Inversely, when there are few infections, DALYs and costs are lower. Moreover, the correlation between DALYs and costs is not perfect, there is large variation in both measures (Figure 5A).

**Figure 5:**
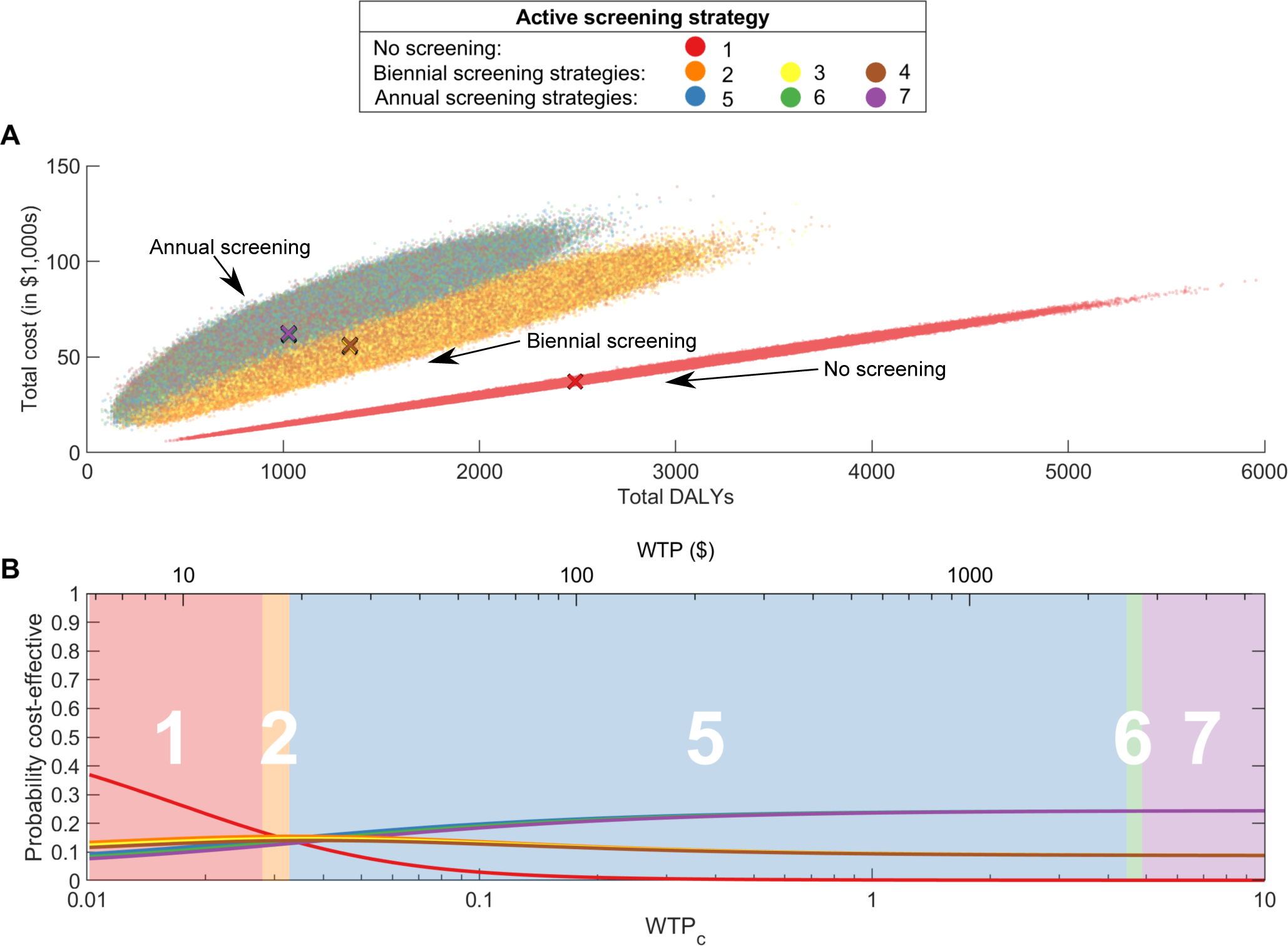
The cost-effectiveness of active screening strategies. (A) Cost-effectiveness plane showing the total cost of a strategy and the associated total number of DALYs. Mean values for each strategy are shown by the coloured crosses. (B) Cost-effectiveness acceptability curves (CEACs) for each strategy are shown by lines, with the cost-effectiveness acceptability frontier (CEAF) shown by the numbered background colour, which demonstrated the values for the ICER. WTP is shown in 2018 USD on the top and as the *WTP*_*c*_ coefficient on the bottom, where the coefficient is the multiplier of the GDP per capita of the DRC. See Table 2 for the full descriptions of the strategies.

There is a similar pattern when active screening exists, but the range of introduced costs and the number of occurring treatments increases, expanding the variance in outcomes and weakening the correlation between costs and DALYs. However, reducing the screening interval both increases the costs and reduces the number of DALYs incurred, all other things held equal. Results for alternative numbers of zero-detections remain marginal over all parameter values (Figure 5A). There are significant differences in the outcomes of strategies with different screening intervals, but we reiterate that the differences are robust to different numbers of active zero-detections (Table 2, Appendix 5 Figure 4).

We have calculated the probability a strategy is cost-effective by taking the proportion of our one million simulations for each strategy that has the largest NMB at each specific value of WTP (Figure 5B), thereby producing cost-effectiveness acceptability curves (CEACs) [51]. The strategy with no active screening has the highest probability of being cost-effective for low WTP, however, the probability is approximately 50% for the lowest WTP (at cost-neutral values), indicating that performing any active screening might still be cost effective. The strategy with no active screening has negligible probability of being the cost-effective (*<* 1%) for approximately *WTP*_*c*_ > 0.23. For the highest WTP values, Strategy 7 (annual screening with three zero-detections), has the highest probability, with similar probabilities for the other strategies with annual screening. Given the probability of being cost-effective for high WTP is so similar for Strategies 5, 6 and 7, it is difficult to make a recommendation about which is better, and so the most cautious strategy (Strategy 7) should likely be favoured, given the minimal change in costs.

It is notable that for certain values of WTP, the strategy with the highest probability of cost-effectiveness does not always correspond with the preferred strategy according to the ICER. These calculations are demonstrated by the cost-effectiveness acceptability frontier, which is shown as the shaded and numbered background (Figure 5B). The CEAF is the optimal strategy according to the maximimum expected NMB (rather than the highest probability of the NMB). Discrepancies between the highest mean NMB and the probability of the highest NMB occur because of asymmetric distributions in our parameters, but it is the CEAF which is used to provide strategy recommendations for a given WTP threshold for risk-neutral decision makers [52].

In the lower-prevalence health zone of Mosango (Appendix 3) we find similar results, with a recommendation for annual screening over biennial screening at moderate WTP values or no active screening at lower WTP values.

## Discussion

To achieve the goal of eliminating gHAT it is useful to have robust models that can inform policy makers about the potential of different intervention strategies [53]. As such, we have followed the five principles of the Neglected Tropical Diseases Modelling Consortium (see Appendix 6), which were proposed to improve the quality of communication between modelers and stakeholders [54]. Furthermore, the addition of economic analysis will further develop the use of this work, as to not only evaluate which strategies are able to decrease in infection in the population, but which are cost-effective.

We have presented a stochastic model for individual villages that demonstrates how active screening should be considered, by determining costs of implementing the screening for different screening coverage levels *c*, screening intervals *t*, and the number of zero-detections observed to stop screening *z*_*a*_ and *z*_*r*_. Unsurprisingly, we find that a big factor in choosing a strategy to implement is how much the programme funder, ministry of health or external donor is willing to pay to avert a DALY. Thus, we present our results across a range of WTP values. We have typically used a WTP value of 0.5 of the GDP per capita of the DRC (*WTP*_*c*_ = 0.5), which is commonly used in the literature [41, 42], but note there may be a higher WTP to achieve the additional aim of gHAT elimination. Using *WTP*_*c*_ = 0.5 we find on average that ideally screening would be done approximately yearly with maximal screening coverage and ceased when a infection is found in a single screening (*c* = 90%, *t* = 0.67 years, *z*_*a*_ = 1) (Figure 3). While the optimum for the screening interval is found to be 0.67 years, if there is a higher proportion of infection eventually treated in the population than the assumed *p*_*t*_ = 27%, the optimal interval is larger (1.25 years for *p*_*t*_ = 100%). Practically these intervals might be difficult to implement, so we believe that the current work supports the implementation of yearly screening. It is noted that screening coverage will rarely be able to be achieved this high and so multiple visits where no infection is observed may be necessary to optimise control, although the model shows no significant differences in cost-effectiveness. This is in line with WHO guidelines of annual active screenings until there have been three consecutive years of no new cases, followed by a further screening with no cases three years after cessation of activities [4].

In particular, we note that while we assumed that reactive screening should immediately resume upon identification of an infection through passive surveillance, the time interval for reactive screening to begin has little effect on the results (see Appendix 4 Figure 3). Therefore, we conclude that practical concerns about the feasibility of reactive screening do not impact our conclusions, as long as reactive surveillance is deployed within two years of finding an new case through passive surveillance.

In fact, the choice of a low *z*_*a*_ has a high probability of triggering reactive screening (>70%), and therefore, we recommend that logistics for reactive screening are put in place (see Appendix 4 Figure 2). The time-horizon of 30 years is sufficient in the village context to capture the dynamics; when we expanded the horizon to 100 years, we found that roughly 99.1% of costs and 99.8% of DALYs are attributable to the first 30 years (see Appendix 5 Figure 1).

As new treatments and active screening modes are introduced, the costs of the model will change, however, the biggest effect is that of the number of DALYs averted, assuming the WTP threshold is set to a reasonable level. Details of the variation in NMB for different screening diagnostics and medical treatments can be found in Appendix 5 Figure 3.

We also note that from the perspective of a single village (and from the perspective of a risk-neutral payer), we do not put any weight on local elimination beyond that captured by expected DALYs averted, favouring an optimal screening strategy that terminates the programme after a single active zero-detection (*z*_*a*_ = 1), rather than repeated zero-detections to ensure elimination. On the other hand, when it is assumed that a village is susceptible to importations of infection, we find that more active zero-detections are required to maximise the NMB (Figure 3F). Other work has shown that least three zero-detections for villages of this size (*N*_*H*_ = 1, 000) [25] to ensure elimination, but it is unclear how much monetary value we should attribute to meeting EOT targets.

Our finding that a single zero-detection is optimal in Figure 3 is particularly notable when the screening coverage *c* is at the maximum (90%). In this case, there is higher confidence that local gHAT elimination has been achieved, as almost all the population is screened and there are no cases left to be detected, and even if infection temporarily persists after this, there is a large probability it will die out due to stochastic fade out [25]. However, a regular 90% coverage is probably unfeasible, and more realistic screening coverage will require more zero-detections to terminate active screening (Figure 4).

We note that there may be also be additional costs in restarting active screening as reactive screening, in particular if regional cessation results in disbanded trained mobile teams. We have not accounted for this in our model, but it may lend support to a higher number of active zero-detections, which lower the probability of reactive screening once routine active surveillance has ceased (see Appendix 4 Figure 2). We cannot make a recommendation for the number of reactive zero-detections as the impact is negligible on the cost-effectiveness (even less so than the active zero-detections) and the effect is completely outweighed by the stochasticity of the infection dynamics (see Appendix 4 Figure 3).

While many other diseases have established procedures for active case finding and evaluated cost-effectiveness (i.e. TB) few have it done for elimination and no studies have evaluated active screening properties with this level of detail. Bessell et al (2018) [10] found that RDTs can be cost-effective. Sutherland et al (2017) [33] took active screening program properties for granted and instead focused on the combination of active screening, passive surveillance, and vector control. Therefore, this is the first cost-effectiveness paper that examines in detail the relative efficiency of active screening strategies. Furthermore, we have provided the tools for the reader to adapt the analysis to their specific chosen costs (https://christopherdavis.shinyapps.io/optimising-ghat-active-screening/).

Future research is warranted to evaluate the specific characteristics of each village, how villages and health zones (or districts) share costs and the impact it makes on relative efficiency. Moreover, the risk of importation, the impact of potential sero-negative skin-infected cases, and the risk of animal reservoirs would have to be further explored in in-depth epidemiological modeling.

In conclusion, we show that when considering different gHAT interventions, active screening is known to be effective in reducing case numbers and hence the infection in the population [6, 7, 8]. Thus, with a limited number of active screening teams and resources for them to carry out their duties, it is important to optimise their activities with the aim of driving towards elimination.

## Methods and Materials

### Mathematical modelling

To capture the effects of different active screening strategies and the underlying infection dynamics on a village population, we use a stochastic compartmental model from Davis et al (2019) [25]. The stochasticity incorporates the chance events involved in infection transmission into the mechanistic model and is better suited than a deterministic model for the case of optimising active screening for a village population. This is because the population, and hence the number of people infected, is small and so, for the pre-elimination setting, extinction of gHAT can be substantially affected by chance events, and the probability of those chance events. A deterministic formulation, which captures average dynamics, would be less suitable to capture the chance events associated with village level extinction of infection.

The model stratifies the human population into two risk classes: low-risk (the majority of the population), which have a lower exposure to the tsetse and participate randomly in active screening; and high-risk (previously estimated as 9.8% in the study health zone of Kwamouth [35]) with a higher exposure to tsetse that also do not participate in active screening. This structure is supported anecdotally with a fraction of the population, typically working males, tending to work by the rivers, which is the habitat of tsetse [55], and being absent for active screening in the villages [47]. Furthermore, previous modelling work indicates that humans have heterogeneous exposure to tsetse [21, 24, 56]. Rock et al (2015) [21] fitted several risk structures for humans and, using the deviance information criterion (DIC), the model with the risk structure given here best matched to data on screening and incidence from the WHO HAT Atlas [49, 50].

The model classifies a person’s infection status as: susceptible *S*_*H*_ ; exposed (or latent) *E*_*H*_ ; Stage 1 infection *I*_1*H*_ ; Stage 2 infection *I*_1*H*_ ; and hospitalised (and temporarily removed) *R*_*H*_. On exposure to the parasite and upon the bite of an infected tsetse, a person will progress through these infection stages, unless detected in active screening and so treated and moved directly to the hospitalised class. Stage 2 infection is defined as the time when trypanosomes have crossed the blood–brain barrier [57]. There is an additional rate defined as the time when people change infection status from Stage 1 infection to hospitalisation to simulate people being treated through passive surveillance.

In addition, the tsetse are explicitly modelled as the proportion of flies that are in the states of: teneral (unfed and more susceptible to infection than fed flies [58]) *S*_*V*_ ; non-teneral yet uninfected *G*_*V*_ ; exposed (or latent) *E*_*V*_ ; and infected *I*_*V*_. The dynamics of the tsetse are modelled with proportions using ordinary differential equations, since the exact number of tsetse that interact with a given population is difficult to determine (although relative tsetse abundance is being mapped in some areas [59]). However, the effective density ratio, the product of the number of tsetse in a population per human and the probability of human infection per single infective bite, can be inferred by model fitting to the WHO HAT Atlas data [49, 50]. A full description of the mathematical equations in the infection model and the parameters used (taken from Crump et al (2020) [35], as the median of the distributions inferred using MCMC methodology and the aggregate annual data from the WHO HAT Atlas in Kwamouth) can be found in Appendix 1.

### Simulating active screening

In villages targeted for active screening, we consider how the infection dynamics are affected by: the screening coverage *c*; the screening interval *t*; active zero-detections *z*_*a*_; reactive zero-detections *z*_*r*_ (see Table 1). This is modelled by taking all combinations of *c, t, z*_*a*_ and *z*_*r*_ for the values *c* = 0, 1, 5, 10, 15, 20, 25, 30, 35, 40, 45, 50, 55, 60, 65, 70, 75, 80, 85, 90, *t* = 0.25, 0.33, 0.5, 0.67, 0.75, 1, 1.25, 1.5, 1.75, 2, 2.5, 3, 3.5, 4, 4.5, 5 years, *z*_*a*_ = 1, 2, 3, 4, 5 screenings, and *z*_*r*_ = 1, 2, 3 screenings. The specified screening procedure is then implemented in the model by using the screening coverage to randomly select a proportion *c* of the population from the low-risk sub-population, which move into the hospitalised class if they are correctly identified as being exposed to the infection after each screening interval. This process is stopped after number of zero-detections equal to *z*_*a*_. However, if the infection has not been eliminated after the active screening has been halted, there is a chance that a new case can be reported by the individual attending a fixed facility to be tested for the infection. The model explicitly incorporates under-reporting of passive case detections, such that only 27% of cases undetected by active screening will be detected by passive surveillance. This parameter was estimated in model fitting, taken as the median of the distribution inferred using MCMC methodology applied to the aggregate annual data from Kwamouth from the WHO HAT Atlas [49, 50]. Thus, we re-start the screening procedure as reactive screening upon identification of these passive cases, stopping again after the given number of consecutive reactive zero-detections, *z*_*r*_.

The WHO aims for high screening coverage in active screenings for villages in gHAT-endemic foci, but just how high that coverage ought to be is not prescribed. Moreover, guidelines and previous modeling work suggest these screenings should continue annually until there have been three consecutive years of no new cases, followed by a further screening after three years if there are no detected cases [4, 25, 60]. Our modelling aims to provide evidence in support of this strategy or recommend how the strategy could be adapted to make more efficient use of resources.

### Economic modelling

To evaluate the NMB, we consider all component costs and the associated health benefits of implementing an active screening strategy compared to passive surveillance alone. However, we do not consider out-of-pocket expenses, therefore framing the model from the perspective of the funder of the programme. Since we are considering the NMB of an active screening strategy, we only consider costs impacted by active screening but we do not consider costs that are fixed across all strategies, such as maintaining a fixed health centre. We note that the total treatment costs will depend on the quantity of active screening carried out and additionally on the proportion of infections that are detected passively. Since we assume 27% of infections that are not found in active screening are detected through passive surveillance, we can make the worst-case assumption that all these reported infections are treated passively, but none of the unreported infections are treated at all (*p*_*t*_ = 27%). Alternatively, we can use a higher value of the parameter *p*_*t*_ to assume more people are treated that are reported in passive surveillance. The parameter *p*_*t*_ takes values in the interval 27–100%, such that the proportion of infections being treated is between only those reported being treated and all infection being treated.

Using the formulation of the NMB given by Equation 1, we then define the constituent parts of the equation as
follows:

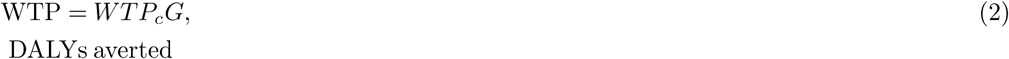

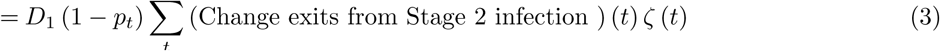

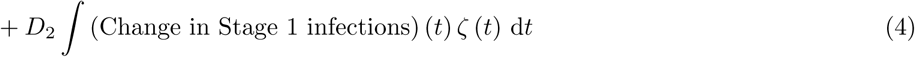

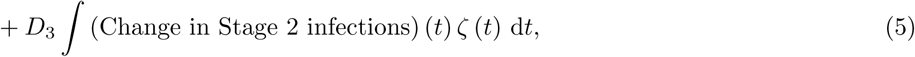

Change in costs

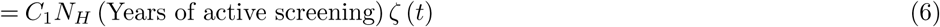

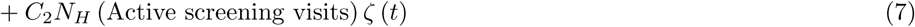

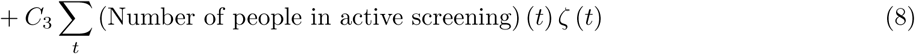

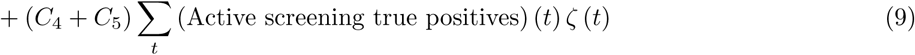

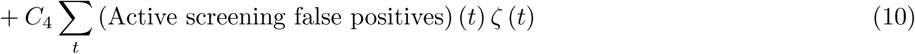

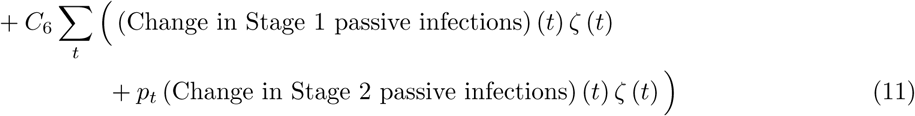

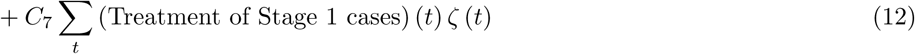

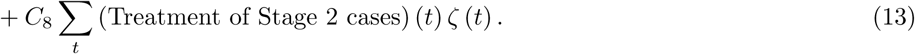

The willingness-to-pay, in 2018 US dollars per DALY, is split into the GDP per capita of the DRC, *G*, and a multiplying factor, *WTP*_*c*_ as is common in the literature [39] (Equation 2). For the health benefit of the intervention, the DALYs averted are the discounted sum of the number of years of life lost (YLL) and years lived with disability (YLD). The number of years of life lost are given by the sum of the change in number of people that exit the Stage 2 infection class multiplied by both the proportion of these infections not treated (1−*p*_*t*_) and the discounted average years of life lost per death (Equation 3). The number of years lived with disability are given by the total time spent in each infection class multiplied by the associated disability weight (Equation 4 and Equation 5).

The change in costs is simply the costs incurred by implementing the active screening strategy and the effect on costs of operating passive surveillance, given as a capital cost of active screening (Equation 6) and the recurrent cost of operating each visit (Equation 7), the number of screening tests carried out (Equation 8), confirmation of the infection and stage determination for the true positive (Equation 9) and negative confirmation for the false positives (Equation 10), the change in testing, confirmation and stage determination for passive infections (Equation 11), and the treatment of detected cases (Equation 12 and Equation 13).

To calculate both the health benefit costs and implementation costs we use an annual discount rate of 3% [43], denoted in the equations by *ζ*(*t*) = exp(−*δt*). The time-horizon used is 30 years, which is sufficient to capture almost all the costs of the active screening programme (see Appendix 5 Figure 1). The cost parameters are fixed values composed as a sum of the cost of the test or treatment, the cost of implementation and the cost of hospitalisation [33]. These costs are also dependent on the type of treatment; we primarily consider active screening to use the CATT algorithm, while Stage 1 and Stage 2 treatments use pentamidine and NECT respectively, but we also consider the additional treatments of fexinidazole and acoziborole. The former we expect to become the standard in 2020 for most gHAT patients [15], and the latter could replace it as a single dose cure a few years later if it passes phase III clinical trials and receives an appropriate recommendation [61] (see Appendix 5 Figure 3). Cost and health parameters are shown in Table 3 with full explanations in Appendix 2.

**Table 3:** Parameters for calculating the NMB. For more detailed calculations of these cost parameters see Appendix 2.

| Parameter | Name | Value |
| --- | --- | --- |
| $WTP_c$ | Willingness-to-pay per DALY | Varies |
| $G$ | GDP per capita for the DRC<sup>1</sup> | \$457.85 |
| $D_1$ | Discounted average years of life lost per death | 21.03 years |
| $D_2$ | Stage 1 disability weight | 0.1330 |
| $D_3$ | Stage 2 disability weight | 0.5432 |
| $C_1$ | Active screening capital cost per person | \$0.22 |
| $C_2$ | Active screening recurrent cost per person | \$0.77 |
| $C_3$ | Active screening test per person <sup>2</sup> | \$1.03 |
| $C_4$ | Confirmation per person <sup>3</sup> | \$10.96 |
| $C_5$ | Stage determination per person <sup>3</sup> | \$26.46 |
| $C_6$ | Passive screening person <sup>3,4</sup> | \$39.05 |
| $C_7$ | Stage 1 treatment per person <sup>5</sup> | \$101.32 |
| $C_8$ | Stage 2 treatment per person <sup>6</sup> | \$783.15 |
| $N_H$ | Population size | Varies |

<sup>1</sup>2018 value used.
<sup>2</sup>We assume the use of the card agglutination test for trypanosomiasis (CATT) test.
<sup>3</sup>We assume confirmation is by microscopy using a blood sample, lymph node aspiration (LNA) and mini Anion Exchange Centrifugation Technique (mAECT).
<sup>4</sup>Passive screening includes the screening test (CATT), outpatient consultation, confirmation and stage determination.
<sup>5</sup>We assume the use of pentimidine.
<sup>6</sup>We assume the use of nifurtimox-eflornithine combination therapy (NECT).

## Data Availability

The authors thank PNLTHA for original data collection and WHO for data access in the framework of the WHO HAT Atlas (Franco et al, 2020)(https://doi.org/10.1371/journal.pntd.0008261).

## Acknowledgments

The authors thank PNLTHA for original data collection and WHO for data access (in the framework of the WHO HAT Atlas [1]). This work was supported by the Bill and Melinda Gates Foundation (www.gatesfoundation.org) in partnership with the Task Force for Global Health through the NTD Modelling Consortium [OPP1184344] (C.N.D., K.S.R. and M.J.K.), the Bill and Melinda Gates Foundation through the Human African Trypanosomiasis Modelling and Economic Predictions for Policy (HAT MEPP) project [OPP1177824] (K.S.R., M.A., and M.J.K.), and EPSRC/MRC via the MathSys Centre for Doctoral Training (C.N.D. and M.J.K.). The funders had no role in study design, data collection and analysis, decision to publish, or preparation of the manuscript.

